# Talking on handsfree and handheld cellphones while driving in association with handheld phone bans

**DOI:** 10.1101/2022.04.08.22273525

**Authors:** Marco H. Benedetti, Li Li, Sijun Shen, Neale Kinnear, M. Kit Delgado, Motao Zhu

**Affiliations:** The Center for Injury Research and Policy, Abigail Wexner Research Institute at Nationwide Children’s Hospital, 700 Children’s Drive, Columbus, OH 43215, USA; Department of Epidemiology and Health Statistics, Xiangya College of Public Health, Central South University, No.932 South Lushan Road, Changsha Hunan 410083 P.R., China; TRL, Crowthorne House, Nine Mile Ride, Wokingham, RG40 3GA, UK; Department of Emergency Medicine, Department of Biostatistics, Epidemiology, and Informatics, and the Penn Injury Science Center, Perelman School of Medicine, University of Pennsylvania, 423 Guardian Dr.Philadelphia, PA 19104, USA; Division of Epidemiology, College of Public Health, The Ohio State University, 1841 Neil Avenue, Columbus, OH 43210, USA; Department of Pediatrics, College of Medicine, The Ohio State University, 370 W. 9^th^ Avenue, Columbus, OH 43210, USA

**Keywords:** Distracted driving, hands-free phone use while driving, handheld phone use while driving, handheld phone policies, traffic safety, policy

## Abstract

**Introduction:** Concurrent use of a cellphone while driving impairs driving abilities, and studies of policy effectiveness in reducing distracted driving have yielded mixed results. Furthermore, few studies have considered how hands-free phone use associates with handheld phone bans. It is not clear whether hand-held phone bans dissuade some drivers from using the phone while driving completely, or whether it simply promotes a shift to hands-free use. The present study estimates the association between handheld phone policies and self-reported talking on handsfree and handheld cellphones while driving.

**Methods:** Our data consisted of 16,067 respondents to annual administrations of the Traffic Safety Culture Index from 2012-2017. Our primary exposure variable was handheld phone policy, and our primary outcome variables were self-reported talking on any phone, self-reported talking on a handheld phone, and self-reported talking on a hands-free phone while driving. We estimated adjusted prevalence ratios of the outcomes associated with handheld phone bans via modified Poisson regression.

**Results:** Drivers in states with handheld bans were 13% less likely to self-report talking on any type of cellphone (handheld or handsfree) while driving. When broken down by cellphone type, drivers in states with handheld bans were 38% less likely to self-report talking on a handheld phone and 10% more likely to self-report talking on a hands-free phone while driving.

**Conclusions:** Handheld phone bans were associated with more self-reported talking on hands-free phones and less talking on handheld phones, consistent with a substitution hypothesis. Handheld bans were also associated with less talking on any phone while driving, supporting a net safety benefit.

**Practical Applications:** In the absence of a national ban on handheld phone use while driving, our study supports state handheld phone bans to deter distracted driving and improve traffic safety.

## 1. INTRODUCTION

Cellphone use while driving negatively affects driving performance.(Caird, Willness, Steel, & Scialfa, 2008; Klauer, et al., 2014; Overton, Rives, Hecht, Shafi, & Gandhi, 2015) The National Highway Traffic Safety Administration (**NHTSA**) estimated that 9.7% of drivers in 2018 were using a cellphone during a typical daylight moment (National Highway Traffic Safety Administration, 2021b). This high prevalence among drivers poses a substantial public health burden; NHTSA estimated that distracted driving contributed to 3,142 deaths in the United States in 2019 (National Highway Traffic Safety Administration, 2021a).

As of July 2021, 24 states have enacted laws that prohibit drivers from holding, talking on, or otherwise operating a handheld cellphone while driving (hereafter referred to as “handheld bans”) to reduce handheld phone use while driving in the United States (Insurance Institute for Highway Safety, 2020; LexisNexis Academic Database, 2020). Literature reviews by McCartt et al. and later Olsson et al. (2020), along with many epidemiological studies of observed and self-reported behaviors, suggest that handheld bans decrease distracted driving (Carpenter & Nguyen, 2015; Ehsani, Ionides, Klauer, Perlus, & Gee, 2016; Li, et al., 2020; Anne T. McCartt, Braver, & Geary, 2003; A. T. McCartt & Geary, 2004; Anne T. McCartt, Hellinga, Strouse, & Farmer, 2010; Anne T. McCartt, Kidd, & Teoh, 2014; Olsson, Pütz, Reitzug, & Humphreys, 2020; Rudisill, Smith, Chu, & Zhu, 2018; Rudisill & Zhu, 2015, 2017; Rudisill, Zhu, & Chu, 2019; Zhu, Rudisill, Heeringa, Swedler, & Redelmeier, 2016). For example, McCartt et al. demonstrated both short- and long-term reductions in observed handheld phone conversations in drivers following handheld bans in New York, Washington DC, and Connecticut (Anne T. McCartt, et al., 2003; A. T. McCartt & Geary, 2004; Anne T. McCartt, et al., 2010). In a study of observed roadside behavior, Rudisill and Zhu (2017) found that universal handheld bans were associated with lower odds of handheld phone conversations across multiple demographic sub-groups and geographic regions (Rudisill & Zhu, 2017). Furthermore, Rudisill et al. (2019) estimated that handheld calling bans were significantly associated with 40% fewer self-reported handheld phone conversations (Rudisill, et al., 2019).

Hands-free calling devices are becoming more widely available, yet the literature on the association between handheld phone bans and hands-free phone use while driving is relatively scarce. A descriptive study by Braitman and McCartt (2010) showed that the percentage of drivers who always use hands-free devices to talk on the phone while driving was higher in states with all-driver handheld phone bans (22%) compared to states with no such bans (13%) (Braitman & McCartt, 2010). Carpenter and Nguyen (2015) utilized a difference-in-differences framework to assess the impact of an all-driver cellphone ban in Ontario, Canada (Carpenter & Nguyen, 2015). The ban was associated with higher hands-free phone use and less handheld and overall phone use while driving, lending support for the hypothesis that handheld bans lead drivers to substitute handheld devices for hands-free ones (hereafter referred to as the “substitution hypothesis”). The descriptive results of Braitman and McCartt are consistent with the substitution hypothesis, however to the authors’ knowledge, an associational study of handheld phone bans and drivers’ self-reported phone behaviors in the United States is not yet available.

To better understand the association between handheld phone policies and self-reported phone use while driving in the United States, we analyzed data from the Traffic Safety Culture Index (**TSCI**), an annual national survey conducted by the AAA Foundation for Traffic Safety. We hypothesized that handheld phone bans were associated with (i) lower rates of self-reported talking on a handheld phone while driving; (ii) higher rates of talking on a hands-free phone while driving; and (iii) a net reduction of self-reported talking on any type of cellphone (either handheld or hands-free) while driving. Taken together, evidence for the first two hypotheses corroborates the substitution hypothesis, which, independent of a net decrease in talking on any phone while driving (i.e., hypothesis (iii)), could be viewed as a form of harm reduction from a public health perspective.

## 2. METHODS

### 2.1 Study Sample

Our study consisted of 16,067 respondents to annual administrations of TSCI for the years 2012-2017. TSCI subjects were drawn from KnowledgePanel, a larger online panel managed by the platform Growth from Knowledge (GfK). Although online panels are prone to selection bias, KnowledgePanel takes steps to mitigate this issue. KnowledgePanel uses address-based sampling from the United States Postal System’s delivery sequence file, initiating contact through non-internet means, and providing non-internet households with notebooks and internet services. In addition, the data include weights that account for differential sampling probability, and which are adjusted via iterative proportional fitting so the weighted sample is representative of the US population with respect to age, gender, race/ethnicity, education, income, household size, and metropolitan status (AAA Foundation for Traffic Safety, 2018). During the study period, the average overall response rate among eligible respondents was 58%, with annual response rates ranging from 55% (2012) to 61% (2017).

We included respondents who indicated they had driven in the last month and aged 21 and older. Because certain handheld phone policies applied only to those under age 21, our age criterion ensured that laws were uniformly applied to all study participants. We only used complete cases, resulting in excluding approximately 8% of the remaining eligible respondents. All data were de-identified and therefore exempt from IRB review at Nationwide Children’s Hospital.

### 2.2 Study Variables

Our outcome variables were binary indicators of (i) self-reported talking on any type of cellphone while driving (either handheld or hands-free), (ii) self-reported talking on a hands-free phone while driving, and (iii) self-reported talking on a handheld phone while driving. Survey respondents were originally presented with Likert-scale responses to questions about these three outcomes. In the 2012-2016 surveys, respondents were first asked how many times they talked on a phone (either handheld or handsfree) while driving in the last month. If a respondent indicated that they talked on a cellphone at least once, the outcome variable for talking on any cellphone was coded as a 1, and 0 otherwise. Respondents who reported talking on any cellphone while driving were also presented with the following question:

*When you talk on your cellphone while driving, do you usually hold the phone in your hand, or do you use a hands-free device?*

1. *I always hold the phone in my hand*
2. *I usually hold the phone in my hand*
3. *I hold the phone in my hand about half the time, and use a hands-free device about half of the time*
4. *I usually use a hands-free device*
5. *I always use a hands-free device*

Unless the respondent reports that they always use a hands-free device (5), we code the outcome variable for talking on a handheld phone as a 1. If the respondent either (i) always uses a handsfree device; or (ii) they have not talked on any type of cellphone while driving in the last 30 days, the response variable for handheld phone use is coded as a 0. The same process was used to generate the outcome variable for talking on hands-free cellphones.

For 2017, TSCI was revised and asks drivers separate questions about talking on handheld phone and handsfree phones, with similar Likert-scale response options. We converted the responses for these question to binary indicators of talking on handheld and hands-free phones, and responses to both questions were used to create binary indicators of any cellphone use for year 2017.

Our primary explanatory variable was handheld phone policy. We assigned policies to respondents by comparing the effective dates of their state cellphone policies to their survey date. We obtained state handheld phone policies from the Insurance Institute for Highway Safety and legal database LexisNexis Academic (Insurance Institute for Highway Safety, 2020; LexisNexis Academic Database, 2020). Two research assistants coded the laws independently, and a group discussion with a third researcher and a lawyer resolved any disagreements. These policy data are provided in a supplemental file.

We coded handheld phone policies into two categories. The category “handheld ban” denoted any policy that prohibited non-emergency operation of a handheld phone, such as talking while holding, reading, or manual typing. The category “no handheld ban” comprised any remaining policies, including but not limited to texting bans, which only prohibited manual typing or reading of text messages, emails, etc., and young driver bans, which did not apply to our study population. Although multiple distinct types of policies were coded as “no handheld ban,” we grouped them because none of them prohibited talking on a handheld phone while driving, one of our three outcome variables. Acknowledging some conceptual differences between policies grouped under “no handheld ban,” we describe in the Supplemental Material an alternative, three-level policy variable, plus descriptive and regression analyses using this categorization (**Section 3 of the Supplemental Material**).

### 2.3 Statistical Analysis

We began our analysis by creating survey-weighted cross-tabulations of the outcomes with demographic and policy variables, along with corresponding 95% confidence intervals. As part of these descriptive analyses, we also performed Rao-Scott χ-square tests to test for between-group heterogeneity in the outcomes.

To infer about the associations between our three outcomes with handheld phone bans, we then fit modified Poisson regression models with robust standard errors (Zou & Donner, 2013), which log-transformed the expected outcomes. Acknowledging possible unmeasured residual correlation between respondents in the same state, we compared the model fit of exchangeable and independent correlation structures for respondents in the same state via quasi-likelihood under independence criteria (*QIC*). An exchangeable correlation structure was superior when modeling talking on a hands-free phone. In contrast, an independent correlation was superior when modeling talking on a handheld phone and talking on any cellphone.

Our regression models controlled for respondent age, gender, household income, education, race/ethnicity, metropolitan status, household size, and census region. These covariates comprised all factors used in sample selection and weighting, thereby improving our ability to infer about the US adult driving population without applying survey weights (Gelman, 2007). We also included year in our model. Because the variables year, age, and household size could have been treated as either numeric and categorical, we selected the combination of numeric and categorical variables that minimized the *QIC*_*u*_, which adds a penalty to the *QIC* based on the number of model parameters. This procedure resulted in all three variables being treated as categorical. The linear component of our models is provided in **Section 2 of the Supplemental Material**.

We fit two models each to the outcomes of talking on a handheld phone while driving and talking on a hands-free phone while driving. The first of these models was fit to data consisting of all drivers, and the second only included drivers who self-reported talking on the phone while driving. For the outcome of talking on any cellphone while driving, we only modeled data consisting of all drivers. A two-sided test of the regression coefficient corresponding to handheld phone bans determined statistical significance. Because our conclusions were based on five adjusted regression coefficients, we applied a Bonferroni-Holm correction to achieve an overall type-I error rate of 0.05. Confidence intervals and *p-*values in descriptive analyses, unadjusted *p*-values, and *p-*values in the supplemental materials were not corrected, as these results were either exploratory or of secondary importance for the present study. All models were fit using SAS Ver. 9.4.

Nearly all respondents excluded due to missing data had complete covariates but were missing at least one outcome variable. Because most imputation methods would preserve the associations between outcomes and covariates, we used complete case analysis.

## 3. RESULTS

Descriptive statistics of the study sample are presented in **Table 1**. Seventy percent of drivers reported talking on any type of cellphone (handheld or handsfree), 50.7% reported talking on a hands-free phone, and 49.0% reported talking on a handheld phone while driving. Higher-income, higher-educated, and younger drivers self-reported talking on hands-free and any cellphone more than their counterparts (*p* < 0.001 for all corresponding Rao-Scott χ-square tests). Younger drivers also self-reported talking on handheld phones while driving more than older drivers (*p* < 0.001). However, except for the lowest income group, there was little apparent income-based difference in self-reported talking on handheld phones while driving. Similarly, drivers with less than a high school education self-reported talking on handheld phones less than drivers with higher education levels (<0.001). Drivers in states with handheld bans self-reported talking on any type of cellphone while driving less than drivers in states without handheld bans (63.9% vs. 72.7%; *p* < 0.001). Drivers in states with handheld bans self-reported talking on a hands-free phone while driving more than drivers in states without handheld bans (56.9% vs. 48.0%; *p* < 0.001). Conversely, drivers in states with handheld bans self-reported talking on a handheld phone while driving less than drivers in states without handheld bans (33.8% vs. 55.8%; *p* < 0.001).

**Table 1:**
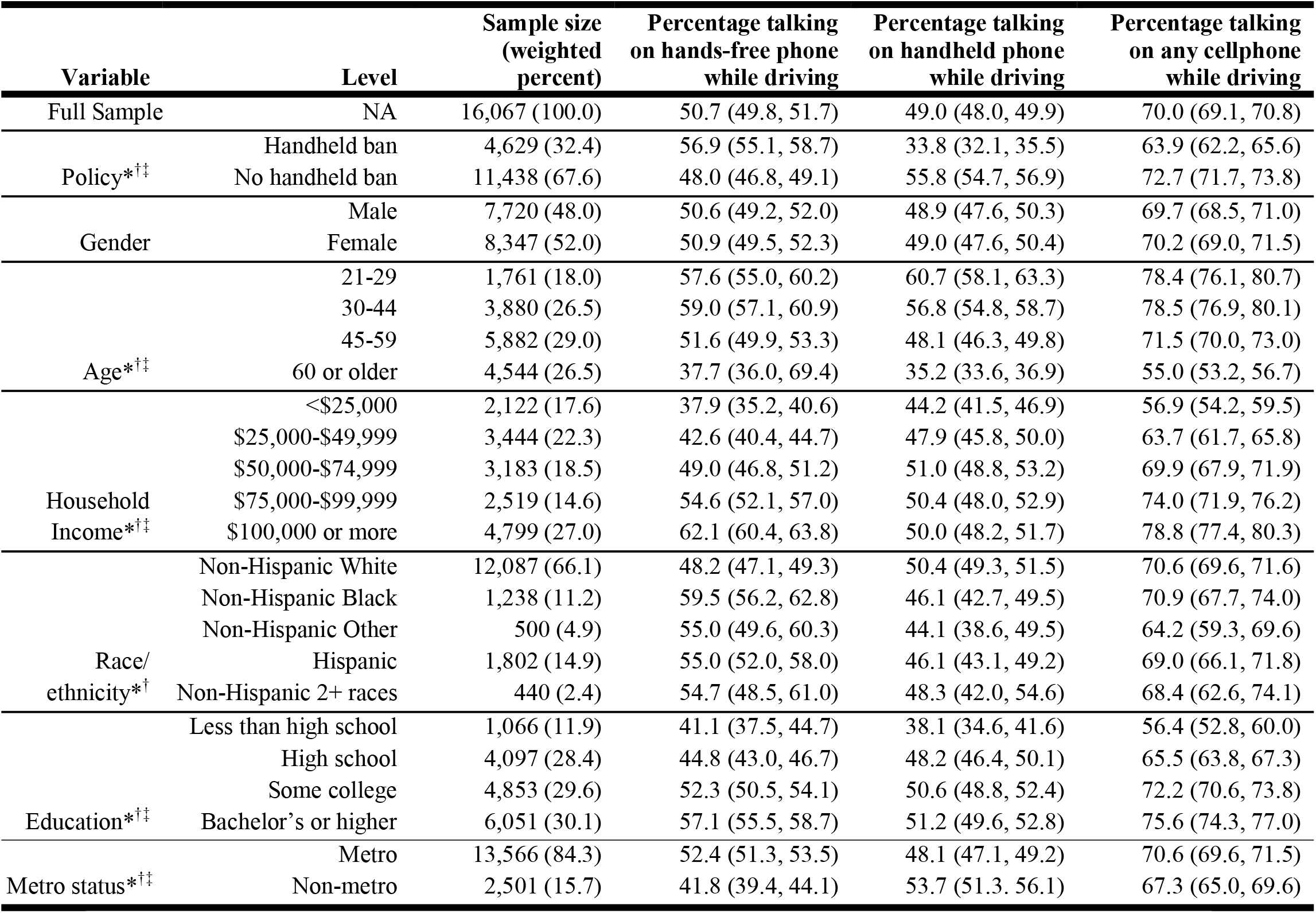
Weighted sample characteristics and percentages (and 95% CIs) of self-reported talking on hands-free, handheld, and any cellphone among drivers in TSCI, years 2012-2017. Superscript symbols in the first column indicate that the *p*-value corresponding to a Rao-Scott χ-square test for between-group heterogeneity in the outcome is less than 0.05. *Handsfree; †handheld; ‡any cellphone (handheld or handsfree)

Survey-weighted cross-tabulations of sociodemographic characteristics and policy status are reported Supplemental Material, along with an assessment of covariate balance between the treatment groups (**Table S1, Section 1 of the Supplemental Material**).

Weighted percentages of driver-reported frequencies of using hands-free and handheld devices when they talk on the phone while driving are presented in **Table 2**. The percentage of drivers who talked on the phone while driving and reported they only spoke on a hands-free phone was more than twice as high in states with handheld bans than in states without them (45.3% vs. 21.8%). In states with handheld bans, only 10.8% of drivers who talk on the phone while driving did so exclusively on handheld devices, compared to 35.5% in states with no handheld bans.

**Table 2:** Weighted percentages (with 95% CIs) of self-reported frequency of handheld and hands-free phone use among drivers who self-reported talking on a cellphone while driving^a^.

| **Policy** | **Always use**<br>**handheld** | **Usually use**<br>**handheld** | **Half handheld**<br>**half hands-free** | **Usually use**<br>**hands-free** | **Always use**<br>**hands-free** |
| --- | --- | --- | --- | --- | --- |
| Handheld ban | 10.8 (9.3, 12.3) | 12.9 (11.2, 14.7) | 9.1 (7.8, 10.6) | 21.8 (19.7, 23.9) | 45.3 (42.8, 47.8) |
| No handheld ban | 35.5 (34.1, 36.9) | 18.5 (17.4, 19.7) | 9.0 (8.1, 9.9) | 15.2 (14.1, 16.3) | 21.8 (22.6, 23.1) |

<sup>a</sup>This question was only asked in the years 2012-2016.

Results from modified Poisson regression models, including unadjusted and adjusted prevalence ratios (**PR**) and corresponding 95% Confidence Intervals (**CI**s) are presented in **Table 3**. After controlling for demographic, economic, and survey-related factors, drivers in states with handheld bans were 13% less likely to self-report talking on any type of cellphone (handheld or hands-free) while driving than drivers in states with no handheld phone ban (PR = 0.87; 95% CI 0.84, 0.91; *p* < 0.001). Drivers in states with handheld bans were also 38% less likely to self-report talking on a handheld phone while driving than drivers in states with no handheld ban (PR = 0.62; 95% CI 0.59, 0.65; *p* < 0.001). Furthermore, drivers in states with handheld bans were 10% more likely to self-report talking on a hands-free phone while driving than drivers in states with no handheld ban (PR = 1.10; 95% CI 1.05, 1.16; *p* < 0.001).

**Table 3:** Unadjusted and adjusted prevalence ratios (handheld ban vs. no handheld ban) and corresponding 95% confidence intervals for modified Poisson regression models applied to three different outcomes.

| Sub-sample<sup>a</sup> | Comparison<sup>b</sup> | Talking on a<br><u>hands-free phone</u><br>while driving<sup>c</sup> | Talking on a<br><u>handheld phone</u><br>while driving<sup>c</sup> | Talking on <u>any</u><br><u>phone</u> while<br>driving<sup>c</sup> |
| --- | --- | --- | --- | --- |
| All Drivers | HH ban vs. none (unadjusted) | 1.17 (1.10, 1.24)*** | 0.59 (0.55, 0.63)*** | 0.86 (0.83, 0.90)*** |
|  | HH ban vs. none (adjusted) | 1.10 (1.05, 1.16)*** | 0.62 (0.59, 0.65)*** | 0.87 (0.84, 0.91)*** |
| Drivers who talk on<br>phone while driving | HH ban vs. none (unadjusted) | 1.36 (1.32, 1.41)*** | 0.68 (0.65, 0.74)*** | -- |
|  | HH ban vs. none (adjusted) | 1.26 (1.21, 1.31)*** | 0.71 (0.68, 0.74)*** | -- |

**a**Analyses were applied to all drivers and only those who talk on the phone while driving.
**b** Adjusted prevalence ratios control for age, gender, income, education, race/ethnicity, metropolitan status, household size, census region, and year.
**c** Asterisks denote the magnitude of the $p$ -values according to the following scale: \*\*\* $p < 0.001$ ; \*\* $0.001 < p < 0.01$ ; \* $0.01 < p < 0.05$ . $p$ -values for the adjusted models were corrected via Bonferroni-Holm. **HH** = Handheld.

Among drivers who talk on the phone while driving, those in states with handheld bans were 26% more likely to self-report talking on a hands-free phone while driving (PR = 1.26; 95% CI 1.21, 1.31; *p* < 0.001), and 29% less likely to self-report talking on a handheld phone while driving (PR = 0.71; 95% CI 0.68, 0.74; *p* < 0.001).

## 4. DISCUSSION

Our study analyzed self-reported talking on hands-free, handheld, and any phone while driving in association with handheld phone policies in the United States. We found that handheld bans were associated with lower self-reported talking on a cellphone while driving. When broken down by phone type, handheld bans were associated with less self-reported talking on a handheld phone while driving and more self-reported talking on a hands-free phone. This pattern was true among all drivers and among drivers who self-reported talking on cellphones while driving, and it corroborates the substitution hypothesis. Our study lends support for all-driver handheld phone bans as a means of reducing talking on phones while driving, while also closing the research gap on hands-free phone use while driving that had previously been addressed by Braitman and McCartt (Braitman & McCartt, 2010) and Carpenter and Nguyen (Carpenter & Nguyen, 2015).

Our descriptive results provided preliminary corroboration of our hypotheses about handheld phone bans and talking on handheld, hands-free, and any cellphone while driving. In addition, several patterns were revealed when our outcomes were cross-tabulated with demographic and economic variables. For instance, there was a clear gradient relationship between income and talking on handsfree phones, yet little apparent relationship between income and talking on handheld phones. It is possible that that this was due to wealthier drivers being able to afford (i) cars that have built-in handsfree capability; and/or (ii) external devices that facilitate handsfree talking. Further study of the relationship between income and handheld/handsfree behavior, in particular understanding if income is a barrier to handsfree phone use, is a promising avenue for future work.

Our results corroborated the substitution hypothesis and demonstrated that handheld phone bans were associated with a reduction in talking on any type of cellphone, either handheld or handsfree. We posit that, independent of a net reduction in talking on any cellphone type, substituting a hands-free phone for a handheld one is preferable from a safety standpoint. Field and simulator studies identify decreased performance resulting from secondary devices distracting visually and/or cognitively while driving (Basacik, Reed, & Robbins, 2011; Lipovac, Đeric, Tešic, Andric, & Maric, 2017; Parkes, Luke, Burns, & Lansdown, 2007; Ramnath, Kinnear, Chowdhury, & Hyatt, 2020). The National Transportation Safety Board (NTSB) advocates for laws that ban all forms of non-emergency personal electronic device use while driving (National Transportation Safety Board, 2020). However, naturalistic driving studies have found that drivers speaking on hands-free phones exhibited little to no increased risk in potentially critical driving events (Gregory M. Fitch, Grove, Hanowski, & Perez, 2014; Gregory M Fitch & Hanowski, 2011; Gregory M Fitch, et al., 2013; Metz, Landau, & Hargutt, 2015). These studies suggest that many of the risks associated with handheld phone use could be attributed to manual and/or visual distractions caused by concurrent sub-tasks, such as dialing, reading or typing (Gregory M. Fitch, et al., 2014; Gregory M Fitch & Hanowski, 2011; Gregory M Fitch, et al., 2013; Metz, et al., 2015). For example, Fitch et al. found that speaking on portable and integrated hands-free devices, in the absence of any other manual or visual distractions, was not associated with a higher risk of safety-critical events (Gregory M Fitch, et al., 2013). Therefore, the current literature supports substituting a handheld phone for a hands-free one as a form of harm reduction. While the risks associated with phone use while driving may be uncertain in some contexts, no phone use while driving is undeniably the safest behavior and should be encouraged by policymakers. Moreover, as in-vehicle communication devices evolve and become more widely available, researchers and policymakers alike should consider how this technology impacts traffic safety and the efficacy of policies like handheld phone bans.

Our finding that handheld phone bans were associated with less talking on handheld phones while driving was consistent with previous studies of both roadside observations (Anne T. McCartt, et al., 2003; A. T. McCartt & Geary, 2004; Anne T. McCartt, et al., 2010; Rudisill & Zhu, 2017; Zhu, et al., 2016) and self-reported behavior (Braitman & McCartt, 2010; Carpenter & Nguyen, 2015; Li, et al., 2020; Rudisill, Smith, et al., 2018; Rudisill, et al., 2019). The reductions in observed and self-reported handheld phone use associated with handheld phone bans from these studies ranged from 24% to 65%. Our estimate of 38% lower self-reported talking on handheld phones while driving fell within this range. We also found that handheld phone bans were associated with less talking on any phone, either hands-free or handheld. This association suggests that, while some substitution may occur, handheld bans are still associated with a net reduction in talking on the phone while driving.

While prior research consistently supports handheld phone bans/restrictions to reduce handheld phone use while driving, corresponding reductions in motor vehicle crash fatalities are less consistent. For example, Lim and Chi did not find that cellphone bans were significantly associated with lower fatal crash rates among all drivers, however they did detect reductions for younger age groups (Lim & Chi, 2013). Other studies have found that handheld bans were significantly associated with reductions in fatalities (more comprehensive reviews are available elsewhere (Anne T. McCartt, Hellinga, & Bratiman, 2006; Anne T. McCartt, et al., 2014; Zhu, et al., 2021)), however it may be argued that some of the detected reductions were of small magnitude (Rudisill, Chu, & Zhu, 2018; Zhu, et al., 2021). While the findings of the present study are encouraging, continued research on the impact of various distracted driving interventions on motor vehicle crashes is necessary.

### 4.1 Limitations

Our study’s first and primary limitation was the data’s narrow temporal scope. Because few states implemented handheld phone bans during the study period, we preferred a cross-sectional analysis of the entire data set rather than a quasi-experimental analysis of a data subset. A quasi-experimental analysis of the impacts of Illinois’ handheld phone ban on self-reported talking on handheld, handsfree, and any cellphones while driving was largely consistent with the present study (Benedetti, et al., 2022). Our second limitation was response bias, which can in any occur study of self-reported behavior. False reporting may have been more prevalent in states with handheld bans due to respondents’ fear of consequences resulting from self-reporting illegal behavior. Although TSCI surveys were anonymized, our results should be interpreted with this caveat in mind. Third, our study excluded younger drivers, who are at high risk of distracted driving and motor vehicle crashes (Buckley, Chapman, & Sheehan, 2014). Li et al. (2020) estimated that concurrent all-driver and young-driver handheld calling bans were associated with 19% lower self-reported talking on any phone while driving among respondents to the Youth Risk Behavior Survey (Li, et al., 2020). Fourth, our study only considered talking on cellphones while driving, and did not consider other actions associated with distracted driving like texting or changing music (Zhu, Rudisill, Rauscher, Davidov, & Feng, 2018). Finally, we acknowledge the possibility that unmeasured factors that were not accounted for in our model may have confounded the associations reported in the present study, which by itself was not capable of identifying a causal relationship between handheld bans and cellphone behavior. Similarly, while our results are consistent with a substitution hypothesis, our data and study design do not allow us to prove such a relationship between state policies and individual behaviors exists.

## 5. Conclusions

Handheld phone bans were associated with higher rates of self-reported talking on hands-free phones while driving and lower rates of self-reported talking on a handheld phone, which was consistent with a substitution hypothesis. Handheld bans’ association with less talking on all phones, particularly handheld ones, supports a net safety benefit.

## 6. Practical Applications

In the absence of prohibiting handheld phone use while driving at the national level, the results of our study support state handheld phone bans as a means of harm prevention and reduction. As of July, 2021, 26 states have yet to enact handheld phone bans, therefore there is still considerable opportunity to improve traffic safety by enacting polices that reduce distracted driving.

## Data Availability

The study's survey data can be obtained by contacting the AAA Foundation for Traffic Safety and submitting a data request form. Programs for data processing and analysis can be obtained by contacting the corresponding author.

## Acknowledgements and Declarations

The authors sincerely thank the AAA Foundation for Traffic Safety, especially Dr. Woon Kim and Leon Villavicencio, for assistance in procuring the TSCI data and for providing valuable feedback on the manuscript.

This research did not receive any specific grant funding from funding agencies in the public, commercial, or not-for-profit sectors.

## Declarations of interest

none

The authors declare no other papers under review or published in another journal that use similar methods or data.

