## Supplemental Materials for "Talking on handsfree and handheld cellphones while driving in association with handheld phone bans"

**Section 1 – Cross-tabulations of Policy Status with Socio-demographic Variables:**

Table S1 presents survey weighted percentages of socio-demographic variables broken up by handheld phone ban policy. The goal of this table is to assess policy-based balance (or lack thereof) in covariates included in our modeling framework.

The distributions of gender and age exhibit balance across policy groups. Drivers in states with handheld bans were more likely to fall into the highest income category. They were less likely to be non-Hispanic white, more likely to be Hispanic, and more likely to live in a metropolitan area, compared to drivers in states with texting only bans. The same is true of drivers in states with no/limited bans. There was subtle policy-based imbalance in education. Drivers in states with handheld bans were more likely to have a bachelor’s degree or higher, and less likely to have only a high school education, than drivers in states with texting bans or no/limited bans.

| Variable | Level | Handheld ban | No handheld ban | Texting ban | No/limited ban |
| --- | --- | --- | --- | --- | --- |
| Gender | Male | 48.9 (47.2, 50.5) | 47.4 (46.3, 48.5) | 47.2 (46.0, 48.4) | 48.0 (45.6, 50.4) |
|  | Female | 51.1 (49.5, 52.8) | 52.6 (51.5, 53.7) | 52.8 (51.6, 54.0) | 52.0 (49.6, 54.4) |
| Age | 21-29 | 16.7 (15.4, 18.0) | 17.0 (16.1, 17.9) | 16.9 (15.9, 18.0) | 17.3 (15.2, 19.3) |
|  | 30-44 | 27.6 (26.0, 29.1) | 26.5 (25.5, 27.5) | 26.7 (25.5, 27.8) | 26.1 (24.0, 28.2) |
|  | 45-59 | 29.3 (27.8, 30.7) | 29.4 (28.4, 30.3) | 29.3 (28.2, 30.4) | 29.7 (27.5, 31.8) |
|  | 60 or older | 26.4 (25.0, 27.8) | 27.1 (26.2, 28.0) | 27.1 (26.1, 28.2) | 27.0 (25.0, 29.0) |
| Income | <$25,000 | 15.3 (14.1, 16.5) | 18.6 (17.7, 19.5) | 17.7 (16.7, 18.7) | 21.7 (19.6, 23.8) |
|  | $25,000-$49,999 | 20.3 (18.9, 21.6) | 23.2 (22.3, 24.1) | 22.4 (21.4, 23.5) | 25.9 (23.8, 27.9) |
|  | $50,000-$74,999 | 16.6 (15.4, 17.8) | 19.5 (18.6, 20.4) | 19.9 (18.9, 20.9) | 17.9 (16.1, 19.7) |
|  | $75,000-$99,999 | 14.7 (13.5, 15.9) | 14.6 (13.8, 15.4) | 14.8 (13.9, 15.7) | 13.8 (12.2, 15.4) |
|  | $100,000 or more | 33.1 (31.6, 34.7) | 24.1 (23.2, 25.0) | 25.1 (24.1, 26.2) | 20.7 (18.9, 22.6) |
| Race/ ethnicity | Non-Hispanic White | 57.1 (55.4, 58.8) | 70.6 (69.5, 71.7) | 74.1 (72.9, 75.3) | 58.7 (56.3, 61.1) |
|  | Non-Hispanic Black | 9.4 (8.3, 10.4) | 12.7 (11.9, 13.5) | 12.9 (12.0, 13.8) | 11.8 (10.1, 13.5) |
|  | Non-Hispanic Other | 9.1 (8.0, 10.1) | 2.8 (2.4, 3.3) | 2.8 (2.3, 3.3) | 2.8 (1.9, 3.8) |
|  | Hispanic | 21.0 (19.5, 22.4) | 12.0 (11.2, 12.8) | 8.4 (7.6, 9.2) | 24.0 (21.8, 26.2) |
|  | Non-Hispanic 2+ races | 3.5 (2.9, 4.1) | 1.9 (1.6, 2.2) | 1.7 (1.4, 2.0) | 2.7 (2.0, 3.4) |
| Education | Less than high school | 12.4 (11.2, 13.7) | 11.7 (10.9, 12.5) | 13.9 (12.0, 15.7) | 11.0 (10.1, 11.9) |
|  | High school | 24.0 (22.6, 25.4) | 30.3 (29.3, 31.3) | 30.2 (28.0, 32.4) | 30.4 (29.2, 31.5) |
|  | Some college | 29.7 (28.2, 31.3) | 29.2 (28.2, 30.2) | 29.7 (27.5, 31.8) | 29.0 (27.9, 30.2) |
|  | Bachelor’s or higher | 33.9 (32.3, 35.4) | 28.8 (27.9, 29.8) | 26.3 (24.3, 28.3) | 29.6 (28.5, 30.7) |
| Metro. Status | Non-metro | 7.0 (6.2, 7.8) | 19.3 (18.4, 20.1) | 20.6 (19.6, 21.6) | 14.9 (13.2, 16.6) |
|  | Metro | 93.0 (92.2, 93.8) | 80.7 (79.9, 81.6) | 79.4 (78.4, 80.4) | 85.1 (83.4, 86.8) |

**Table S1:** Survey-weighted percentage of respondents in socio-demographic categories based on policy variables with 95% confidence intervals.

**Section 2 – Modeling Framework**

Here, we present the linear component of our modeling framework for binary policy variables. Interested readers may refer to Zou and Allen for details on fitting Poisson regression models for correlated binary data. For easy of notation, we index observations using only a single subscript. Let $Y_{i}$ denote one of the three self-reported outcome variables for subject $i, i=1,\ldots,n$, let $E\left( \cdot\right)$ denote expectation, and let $I\left( x \right)$ denote an indicator function that equals 1 when $x$ is true, and 0 otherwise.

| $\log\left( E\left( Y_{i} \right) \right)$ | $=$ | $\beta_{0}+ \beta_{1}I\left( Law_{i}=Handheld Ban \right)+\beta_{2}I\left( Gender_{i}=Male \right)+$ |
| --- | --- | --- |
|  |  | $\beta_{3}I\left( Age_{i}=30-44 \right)+\beta_{4}I\left( Age_{i}=45-59 \right)+\beta_{5}I\left( Age_{i}\geq60 \right)+$ |
|  |  | $\beta_{6}I\left( Inc_{i}=\$25,000-\$49,999 \right)+ \beta_{7}I\left( Inc_{i}=\$50,000-\$74,999 \right)+$ |
|  |  | $\beta_{8}I\left( Inc_{i}=\$75,000-\$99,999 \right)+ \beta_{9}I\left( Inc_{i}>\$100,000 \right)+$ |
|  |  | $\beta_{10}I\left( RaceEth_{i}=NH Black \right)+ \beta_{11}I\left( RaceEth_{i}=NH Other \right)+$ |
|  |  | $\beta_{12}I\left( RaceEth_{i}=NH Multiracial \right)+ \beta_{13}I\left( RaceEth_{i}=Hispanic \right)+$ |
|  |  | $\beta_{14}I\left( Ed_{i}= Less than HS \right)+\beta_{15}I\left( Ed_{i}=HS \right)+$ |
|  |  | $\beta_{16}I\left( Ed_{i} =Some College \right)+\beta_{17}I\left( Ed_{i}=Bachelor^{'}s or higher \right)+$ |
|  |  | $\beta_{18}I\left( HH Size_{i}=1 \right)+\beta_{19}I\left( HH Size_{i}=2 \right)+\beta_{20}I\left( HH Size_{i}=3 \right)+\beta_{21}I\left( HH Size_{i}\geq4 \right)+$ |
|  |  | $\beta_{22}I\left( Region_{i}=Midwest \right)+\beta_{23}I\left( Region_{i}=Northeast \right)+\beta_{24}I\left( Region_{i}=South \right)+$ |
|  |  | $\beta_{25}I\left( Metro Status_{i}=Metro \right)+\beta_{26}I\left( Year_{i}=2013 \right)+\beta_{27}I\left( Year_{i}=2014 \right)+$ |
|  |  | $\beta_{28}I\left( Year_{i}=2015 \right)+\beta_{29}I\left( Year_{i}=2016 \right)+\beta_{30}I(Year_{i}=2017)$ |

**Section 3 – Analyses with Three-level Policy Variable:**

Initially when categorizing handheld phone policies, we identified three distinct categories of policies that were relevant to the present study: (i) “handheld bans” comprised policies that prohibit all drivers from holding or operating a handheld device or cellular phone while operating a motor vehicle; (ii) “texting bans” only prohibited all drivers manually typing on or reading a handheld device or cellular phone while operating a motor vehicle; and (iii) “no/limited bans” provided either no restriction on handheld phone use while driving, or stipulated restrictions that did not apply to our study population (for example, they only banned cellphone use among teen drivers). All policies that fell under the category of handheld bans also banned texting while driving. While policies that banned only handheld calling have existed in the past, none were present during the scope of our study. Some policies coded as “handheld bans” included bans on data use while driving whereas others did not, however we grouped them into a single category.

Table S2 provides a cross-tabulation of our outcome variables and a three-level policy. For each of the outcomes, drivers in states with texting bans are very similar to those in states with no/limited bans. Drivers in states with handheld bans self-reported talking on handheld and any cellphones while driving less than their counterparts in states with texting bans and no/limited bans. Conversely, drivers in states with handheld bans self-reported talking on handsfree phones more than those in states with texting bans and no/limited bans.

Table S3 presents a cross-tabulation of the relative frequency at which drivers use handsfree and handheld devices when they talk on the phone while driving, broken up by the three-level policy variable. The percentage of drivers who talked on the phone while driving and reported that they always did so on a handsfree phone higher in states with handheld bans (45.3%) than in states with texting bans (20.8%) and no/limited bans (24.6%). In states with handheld bans, only 10.8% of drivers who talk on the phone while driving did so exclusively on handheld devices, compared to 36.0% in states with texting bans, and 34.1% in states with no/limited bans.

Table S4 presents results from our models with three-level policy variables. We found little evidence that drivers in states with handheld bans were more likely to self-report talking on a handsfree phone than drivers in states with no or limited bans (PR = 1.06; 95% CI = 0.99, 1.14), although the latter category had a small sample size, yielding greater uncertainty. Drivers in states with handheld bans were more likely to self-report talking on a handsfree device than drivers in states with texting only bans (PR = 1.12; 95% CI 1.06, 1.18). Furthermore, drivers in states with no or limited bans were marginally less likely to self-report talking on a handheld phone while driving than those in states with texting only bans (PR = 0.92; 95% CI 0.86, 1.00). An explanation for this association is that drivers in states with no or limited bans may be more prone to communicate via text message while driving, whereas texting only bans lead drivers to favor talking on a handheld phone over texting. However, this explanation is mostly conjecture and based on a weak observed association, hence further research would be needed to test it.

Table S5 presents additional results in which we fit modified Poisson regression models to the outcomes talking on handsfree and handheld phones while driving, however we restrict the analysis to those who self-reported talking on the phone while driving.

| Variable | Level | Percent of total sample | Percentage who self-reported talking on handsfree phone while driving | Percentage who self-reported talking on handheld phone while driving | Percentage who self-reported talking on phone while driving |
| --- | --- | --- | --- | --- | --- |
| Policy  (3-levels) | Handheld ban | 32.4 (31.6, 33.1) | 56.9 (55.1, 58.7) | 33.8 (32.1, 35.5) | 63.9 (62.2, 65.6) |
|  | Texting ban | 52.2 (51.4, 53.0) | 48.0 (46.7, 49.3) | 56.4 (55.1, 57.7) | 73.0 (71.8, 74.1) |
|  | No/limited ban | 15.4 (14.8, 16.0) | 47.8 (45.3, 50.3) | 53.6 (51.4, 56.1) | 72.0 (69.8, 74.2) |

**Table S2:** Weighted sample characteristics and percentages (and 95% CIs) of self-reported talking on handsfree, handheld, and any cellphone among drivers broken up by three-level policy variable.

| Policy | Always use handheld | Usually use handheld | Half handheld half handsfree | Usually use handsfree | Always use handsfree |
| --- | --- | --- | --- | --- | --- |
| Handheld ban | 10.8 (9.3, 12.3) | 12.9 (11.2, 14.7) | 9.1 (7.8, 10.6) | 21.8 (19.7, 23.9) | 45.3 (42.8, 47.8) |
| Texting ban | 36.0 (34.3, 37.7) | 19.1 (17.7, 20.4) | 8.9 (7.9, 9.8) | 15.3 (14.0, 16.5) | 20.8 (19.4, 22.3) |
| No/ limited ban | 34.1 (31.2, 37.0) | 17.1 (15.7, 19.4) | 9.4 (7.5, 11.2) | 14.9 (12.7, 17.1) | 24.6 (22.0, 27.3) |

**Table S3:** Weighted percentages (with 95% CIs) of self-reported frequency of handheld and handsfree phone use among drivers who self-reported talking on a cellphone while driving, broken up by three-level policy variable. This question was only asked in years 2012-2016.

| Models | Policy Comparison | Self-reported talking on a **handsfree phone** while driving | Self-reported talking on a **handheld phone** while driving | Self-reported talking on **any phone** while driving |
| --- | --- | --- | --- | --- |
| Unadjusted | Handheld ban vs. texting ban | 1.14 (1.08, 1.22)*** | 0.58 (0.54, 0.62)*** | 0.86 (0.82, 0.91)*** |
|  | No or limited ban vs texting ban. | 0.89 (0.80, 0.99)* | 0.94 (0.87, 1.02) | 0.99 (0.86, 1.03) |
|  | Handheld ban vs no/ limited ban | 1.29 (1.15, 1.44)*** | 0.61 (0.57, 0.67)*** | 0.87 (0.83, 0.91)*** |
| Adjusted | Handheld ban vs. texting ban | 1.12 (1.06, 1.18)*** | 0.61 (0.57, 0.64)*** | 0.87 (0.84, 0.90)*** |
|  | No or limited ban vs texting ban. | 1.05 (0.98, 1.13) | 0.92 (0.86, 1.00)* | 1.01 (0.97, 1.05) |
|  | Handheld ban vs no/limited ban | 1.06 (0.99, 1.14) | 0.66 (0.61, 0.71)*** | 0.86 (0.83, 0.91)*** |

**Table S4:** Unadjusted and adjusted prevalence ratios and corresponding 95% confidence intervals for modified Poisson regression models applied to three different outcomes with 3-level policy variable. prevalence ratios are adjusted for age, gender, income, education, race/ethnicity, metropolitan status, household size, census region, and year. Asterisks denote the magnitude of the *p*-values according to the following scale: ****p* < 0.001; **0.001 < *p* < 0.01; *0.01 < *p* < 0.05.

| **Models** | **Policy Comparison** | Self-reported talking on a **handsfree phone** while driving | Self-reported talking on a **handheld phone** while driving |
| --- | --- | --- | --- |
| Unadjusted | Handheld ban vs. texting ban | 1.34 (1.30, 1.39)*** | 0.67 (0.64, 0.70)*** |
|  | No or limited ban vs texting ban. | 0.90 (0.82, 1.00) | 0.95 (0.90, 1.01) |
|  | Handheld ban vs no or limited ban | 1.48 (1.34, 1.63)*** | 0.90 (0.66, 0.75)*** |
| Adjusted | Handheld ban vs. texting ban | 1.27 (1.22, 1.33)*** | 0.70 (0.67, 0.73)*** |
|  | No or limited ban vs texting ban. | 1.03 (0.97, 1.10) | 0.92 (0.87, 0.96)*** |
|  | Handheld ban vs no or limited ban | 1.23 (1.15, 1.31)*** | 0.76 (0.72, 0.81)*** |

**Table S5:** Unadjusted and adjusted prevalence ratios and corresponding 95% confidence intervals for modified Poisson regression models applied to two different outcomes with 2- and 3-level policy variables, only among those who self-reported talking on a cellphone while driving. Prevalence ratios are adjusted for age, gender, income, education, race/ethnicity, metropolitan status, household size, census region, and year. Asterisks denote the magnitude of the *p*-values according to the following scale: ****p* < 0.001; **0.001 < *p* < 0.01; *0.01 < *p* < 0.05.
